# Ultrasonic Texture Analysis for Acute Myocardial Infarction Risk Stratification: A Pilot Study

**DOI:** 10.1101/2024.03.26.24304839

**Authors:** Quincy A. Hathaway, Ankush D. Jamthikar, Bernard R. Chaitman, Jeffery Carson, Naveena Yanamala, Partho P. Sengupta

## Abstract

**Background:** Current risk stratification tools for acute myocardial infarction (AMI) have limitations, particularly in predicting mortality. This study utilizes cardiac ultrasound radiomics (i.e., ultrasomics) to risk stratify AMI patients when predicting all-cause mortality.

**Methods:** The study included 197 patients: a) retrospective internal cohort (n=155) of non-ST-elevation myocardial infarction (n=63) and ST-elevation myocardial infarction (n=92) patients, and b) external cohort from the multicenter Door-To-Unload in ST-segment–elevation myocardial infarction [DTU-STEMI] Pilot Trial (n=42). Echocardiography images of apical 2, 3, and 4-chamber were processed through an automated deep-learning pipeline to extract ultrasomic features. Unsupervised machine learning (topological data analysis) generated AMI clusters followed by a supervised classifier to generate individual predicted probabilities. Validation included assessing the incremental value of predicted probabilities over the Global Registry of Acute Coronary Events (GRACE) risk score 2.0 to predict 1-year all-cause mortality in the internal cohort and infarct size in the external cohort.

**Results:** Three phenogroups were identified: Cluster A (high-risk), Cluster B (intermediate-risk), and Cluster C (low-risk). Cluster A patients had decreased LV ejection fraction (P=0.004) and global longitudinal strain (P=0.027) and increased mortality at 1-year (log rank P=0.049). Ultrasomics features alone (C-Index: 0.74 vs. 0.70, P=0.039) and combined with global longitudinal strain (C-Index: 0.81 vs. 0.70, P<0.001) increased prediction of mortality beyond the GRACE 2.0 score. In the DTU-STEMI clinical trial, Cluster A was associated with larger infarcts size (>10% LV mass, P=0.003), compared to remaining clusters.

**Conclusions:** Ultrasomics-based phenogroup clustering, augmented by TDA and supervised machine learning, provides a novel approach for AMI risk stratification.

## Introduction

Globally, acute myocardial infarction (AMI) affects nearly 10% of people over 60 years of age (1). In the United States, the total annual cost of AMI was $85 billion in 2016, with an estimated $40 billion lost due to premature mortality in the preceding decade (2). Unfortunately, despite the success of intervention and evolving guideline-directed treatment, AMI patients continue to have high morbidity and mortality (3). Currently, clinicians use validated risk stratification scoring systems, such as the Global Registry of Acute Coronary Events (GRACE) (4,5) and more recently the GRACE 2.0 score (6), to predict the 6-month and 1-year risk of all-cause mortality following AMI. While guidelines have recommended using the GRACE score as the most robust model for all acute coronary syndrome types (7–9), these scores were developed using clinical trial data long before percutaneous interventions became routine. Moreover, GRACE uses conventional statistical approaches (i.e., logistic regression) with fixed linear assumptions on data behavior and limited variables, resulting in modest discrimination (e.g., C-statistic range for predicting mortality:0.65-0.8) (5,9).

Artificial intelligence (AI) techniques have led to the development of novel methods that includes subjecting images and other inputs to sophisticated algorithms to capture complexity of human health and disease at the level of the individual (10). These methods have achieved remarkable success, especially in disease classification and risk assessments, in several image-based disciplines, such as dermatology, gastroenterology, ophthalmology, oncology, and neuroradiology (10–16), including the development of ‘omics’-based decision support tools (17–21). The application of radiomics to cardiac ultrasound (i.e., ultrasomics), may aid in risk stratification of patients experiencing an AMI by extracting texture-based information from the myocardium. Moreover, the development of automated tools that integrate ultrasomics for AMI risk stratification addresses the existing gap in current guidelines which do not currently integrate cardiac imaging-based information in existing tools like GRACE 2.0 for estimating risk.

In the present study, we used a cluster-then-predict approach for AMI risk stratification. We subjected cardiac ultrasomics information to topological data analysis (TDA)—a robust method to create compressed representations of highly dimensional data to create unique patient phenogroups (22). We illustrate that the ultrasomics phenogroups can provide independent and incremental information to conventional tools like GRACE 2.0 for augmenting 1-year mortality prediction in AMI patients. Moreover, TDA can be effectively combined with machine learning and explainable AI techniques. Accordingly, we also illustrate the ability to develop robust supervised machine-learning algorithms on clustered patients, which can be applied to external data for phenogroup prediction. Since infract size is strongly associated with all-cause mortality in AMI (23), we used the Door-To-Unload in STEMI (DTU-STEMI) Pilot Trial (24) as an external, prospective, multicenter clinical trial cohort to illustrate that the high-risk phenogroup had larger infarct size as observed on cardiac magnetic resonance (CMR) imaging.

## Materials and Methods

### Study Population

For the internal validation dataset, we identified 155 AMI patients retrospectively from electronic medical record of Robert Wood Johnson University Hospital who were admitted over a 6-month period between January 2023 to July 2023. The Institutional Review Board (IRB) of Robert Wood Johnson University Hospital gave ethical approval for this work (#Pro2023001660).

This included 87 patients classified as having a NSTEMI (non-ST-elevation myocardial infarction) and 121 as having a STEMI (ST-elevation myocardial infarction). STEMI was classified per the Joint ESC/ACCF/AHA/WHF Task Force (25). Briefly, this included ECG changes revealing 1) new ST-segment elevation in 2 contiguous leads with greater than 0.1 mV in all leads, with the exception of V2 or V3, 2) new ST-segment elevation in leads V2-V3 greater than 0.2 mV (men > 40 years old), 0.25 mV (men < 40 years old), or 0.15 mV (women), 3) Pre-existing left bundle branch block were further evaluated using the Sgarbossa’s criteria (26,27). Exclusion criteria included (1) patients discharged to institutionalized care, (2) type 2-5 acute myocardial infarction (AMI), (3) co-existing terminal illness such as cancer, (4) alternative diagnosis for elevated cardiac troponin values (e.g. myocarditis, pericarditis, non-ischemic cardiomyopathies, moderate-severe valvular heart disease), (5) pregnancy, and (6) technically insufficient imaging for 2 of the following 3 views: apical 4 chamber (A4C), apical 3 chamber (A3C), and apical 2 chamber (A2C). Of the 208 patients initially enrolled, 53 patients were accordingly excluded from analysis, this included patients with NSTEMI (n=24) and STEMI (n=29). We assessed the performance of the GRACE 2.0 score (6) with the primary outcome of all-cause mortality at one year.

For the external validation dataset, participants were recruited from a prospective, multicenter, randomized DTU-STEMI pilot trial (24). We included 42 participants with CMR data in the current study. Briefly, patients were included in the original randomized pilot trial if they 1) were between 21 and 80 years of age and 2) presented with 1-6 hours of chest pain with documented ST-segment elevation of ≥2 mm in ≥2 contiguous anterior leads or ≥4 mm total ST-segment deviation sum in the anterior leads. Patients were excluded if they had prior AMI, coronary artery bypass grafting surgery, out-of-hospital cardiac arrest requiring cardiopulmonary resuscitation, cardiogenic shock, inability to undergo Impella CP insertion, fibrinolysis within 72 hours of presentation, or contraindications to CMR imaging (24).

For the external validation study infarct size on CMR was used as the primary end point. CMR-quantified infarct size was categorized as large (LGE mass accounting for >10% of the total LV mass) or small (LGE mass accounts for ≤10% of the total LV mass) (28,29). The details of the CMR protocol have been previously described (24). Briefly, patients in the DTU-STEMI trial underwent standard CMR with steady-state free-precession sequence for LV ejection fraction, volumes, and mass analysis on days 3 to 5 and again on day 30 (±7 days). Delayed-enhancement imaging was performed using a 2-dimensional segmented inversion-recovery sequence, 10 minutes after administration of routine extracellular gadolinium contrast (0.15 mmol/kg of body weight). Infarct size was expressed as a percentage of total LV mass. A central core laboratory (Duke Cardiovascular Magnetic Resonance Center, Durham, NC) qualified participating sites, performed quality assessment on the images during the conduct of the study, and manually performed assessment of CMR parameters on deidentified images without knowledge or access to treatment assignment or clinical outcomes. For the external cohort, institutional review boards at each site approved the trial, and patients provided written, informed consent. The study was approved by the Food and Drug Administration (NCT03000270).

### Echocardiography Image Acquisition, Preprocessing, and Semantic Segmentation

Echocardiograms from A4C, A3C, and A2C were utilized in the present studies for both the internal and external validation data analysis. Patients and participants required at least two of the three views to be present to be included in the current study (*see Materials and Methods, section Study Population*). 2D echocardiograms were preprocessed from video formats to DICOM using Sante DICOM Viewer Pro (SanteSoft, Nicosia, Cyprus, Greece). DICOM files containing doppler data, dual ultrasound regions, or other with limited technical views were discarded. A4C, A3C, and A2C multi-beat echocardiogram DICOM files were manually selected. Using echocv (30) (i.e., a semantic segmentation algorithm that automatically defines regions of the heart in echocardiography images through convolutional neural networks (CNNs)) were segmented the region of the left ventricle (LV) in the A4C, A3C, and A2C views.

Compared to the published version of the algorithm, we modified echocv to be executed using Python 3.2 and leveraged TensorFlow 1.15.0 with GPU support, alongside CUDA 10.0. The segmented images were also uniformly resized to a fixed shape of 1024 by 1024 to ensure consistency across various image sources. Otherwise the use of algorithm and its validation has previously been published, specifically for predicting LV remodeling in parasternal long axis echocardiograms (31).Using the semantic segmentation algorithm, a binary mask representing the region of interest (ROI) within the A4C, A3C, and A2C views was achieved (**Figure S1A**). The ROI for each of the three views was then processed to obtained radiomics/ultrasomics-based information.

### Texture-based Feature Extraction

Echocardiography ultrasomics were extracted in Python (v3.7.13) using pyradiomics (v3.0.1) (32), SimpleITK (v2.2.0) (33), pywavelets (v1.3.0), and numpy (v1.21.5) for both the internal and external validation sets. We have previously published using this methodology on the LV (31). Briefly, feature extraction was performed for the 2D ROI using featureextractor() from pyradiomics. Default parameters for extraction, binwidth, resampled pixel spacing, interpolator, label definition, were applied. In total, first-order (n=18), shape (n=9), and texture-based (n=73) features were extracted for each of the echocardiography views (i.e., A4C, A3C, and A2C) (**Figure S1B**).

### Topological Data Analysis (TDA)

The online tool TDAView (34) was used for phenogroup cluster of AMI patients in the internal validation set. TDAView utilizes the Mapper algorithm for TDA. A 1D Mapper filter was applied using Euclidean distance. Number of intervals was defined as 10, with 5 bins. To reduce the overlap between clusters, a 5% overlap was defined. The number of clusters was not fixed. Based on the parameters used in TDAView, three clusters were generated, labeled as Cluster A (n=62), B (n=43), and C (n=50).

### Supervised Machine Learning Classifier

BigML (https://bigml.com. BigML, Inc. Corvallis, Oregon, USA) was utilized for supervised machine learning and to develop a classifier for prediction of patients in Cluster A, B, and C. Weights were applied to Cluster A (weight=1), Cluster B (weight=1.189), and Cluster C (weight=1.023) to address class imbalance. Through the OptiML application (i.e., a supervised machine learning algorithm that compares generated ensembles, deep neural networks, and logistic regression algorithms) 10-fold cross validation was performed and prediction of Cluster A, B, and C phenogroups was performed using only ultrasomics features. Once the model was developed, batch prediction was performed on the external validation set (n=42 participants) to assign phenogroup information.

### Data Availability

All code is made freely available on our GitHub repository https://github.com/qahathaway/AMI_Phenogroups. All data is available by reasonable request.

### Statistics

GraphPad Prism (v10.1.1) and R (v4.1.0) were used for statistical analyses. The Shapiro-Wilk test assessed normality. In normally distributed data with continuous variables, a two-sided Student’s t-test was applied. In non-Gaussian distributed data, the Mann-Whitney test was used. When assessing more than one group of continuous variables, a one-way analysis of variance (ANOVA) was applied. A Dunnett’s multiple comparisons test was used for multiple comparisons in the one-way ANOVA. When assessing more than one group of categorical variables, a non-parametric Kruskal-Wallis test was applied with multiple comparisons testing.

Receiver operating characteristics (ROC) area under the curve (AUC) was created using the BigML platform, utilizing 10-fold cross validation. A Kaplan-Meier curve was generated using the R packages survival (v3.4-0) (35) and survminer (v0.4.9). Stratification of events, assessed as patients at risk for mortality at one year, was performed over 50-day increments for patients in Cluster A, Cluster B, and Cluster C. The *P*-value was calculated using the log-rank test in R. Using the survival package, a Cox Proportional Hazard model (CoxPH) for time-to-event analyses of mortality at one year was assessed. A risk score was generated with the A) GRACE 2.0 score alone, B) GRACE + Cluster A, C) GRACE + LV global longitudinal strain, and D) using all three variables through CoxPH regression. A probability score (i.e., ranging from 0 to 1) for predicting outcomes was generated using the predictRisk function of the riskRegression (v2022.11.28) package in R. The concordance index (C-statistic) was calculated using the pec (v2022.05.04) package in R (36).

## Results

### Study Overview

The current study utilizes an internal validation group of acute myocardial infarction (AMI) patients (n=155) presenting with non-ST-elevation myocardial infarction (NSTEMI) and ST-elevation myocardial infarction (STEMI). Apical 4-chamber (A4C), apical 3-chamber (A3C), and apical 2-chamber (A2C) views were utilized (**Figure 2A**). Using echocardiography-derived ultrasomics, phenogroups were labeled through TDA and applied to the prediction of clinical outcomes, such as time-to-event mortality (**Figure 2B**). A supervised machine learning algorithm was further used to characterize which radiomics features are important in prediction of the phenogroups and generation of risk prediction score. We then evaluated the incremental value of the phenogroups using the internal validation group and explored how assigned phenogroup labels contributed to predicting CMR findings in the external validation group (**Figure 2C**).

### Patient Demographics and Functional Parameters – Internal Validation

Demographic features for patients in the internal validation study presenting with NSTEMI (n=63) and STEMI (n=92) were assessed (**Table 1**). Patients presenting with STEMI were more likely to have a history of congestive heart failure (CHF) (20.65% vs. 1.59%, P=0.0004) and higher Global Registry of Acute Coronary Events (GRACE) Score (120.63 vs. 107.92, P=0.0184), compared to NSTEMI patients, respectively. Patients presenting with NSTEMI were more likely to have a history of coronary artery disease (CAD) (52.38% vs. 19.57%, P<0.0001), chonic kindey disease (CKD) (23.81% vs. 10.87%, P=0.0315), and stroke (17.46% vs. 6.52%, P=0.0324), compared to STEMI patients, respectively. When comparing the groups based on type of AMI, there were no differences in outcomes, including major adverse cardiac events (MACE) at 30 days (P=0.3803), cardiovascular death at 1 year (P=0.8910), and all-cause mortality at 1 year (0.9502).

**Table 1:**
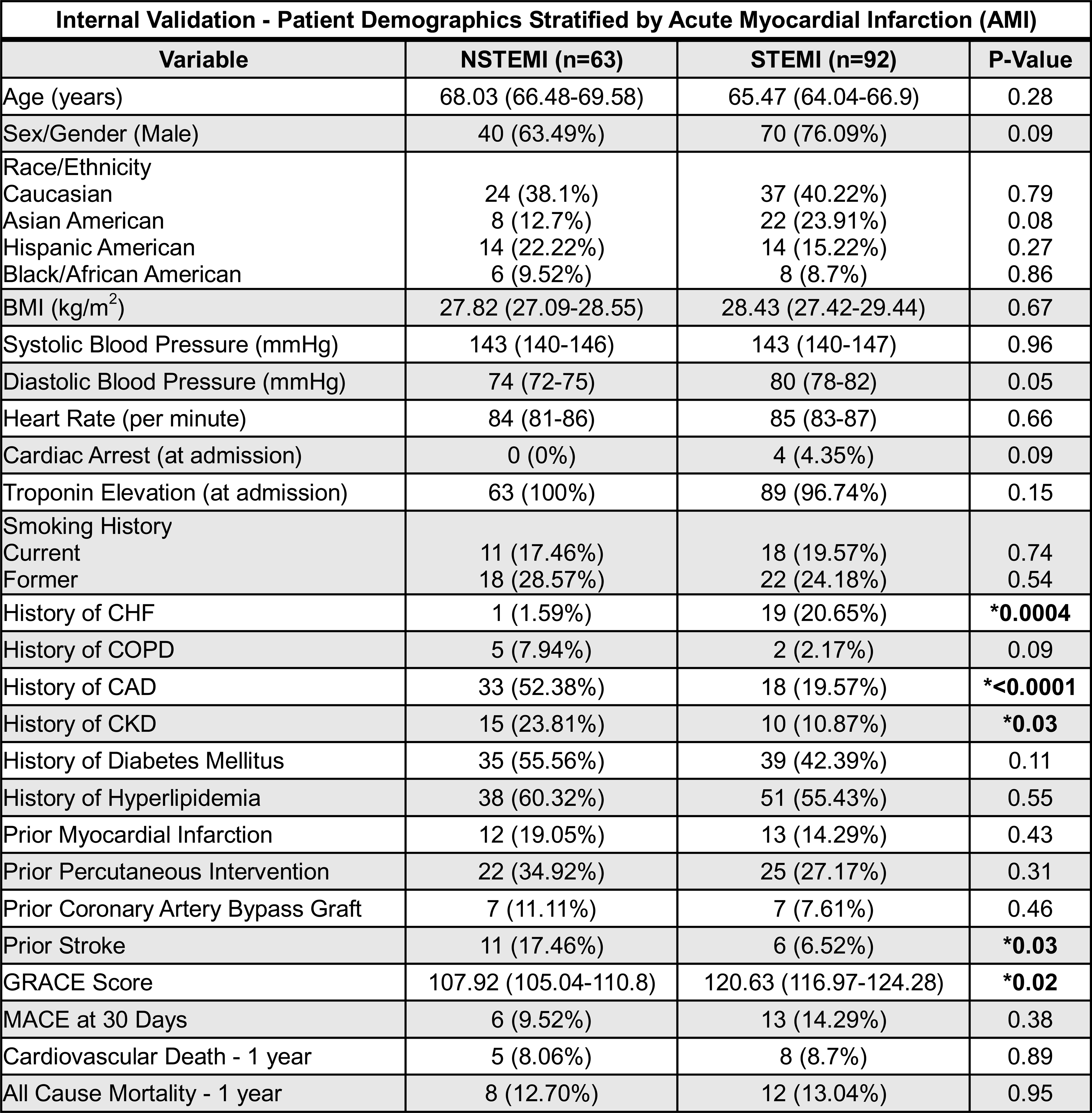
Patient Demographics of the Internal Validation Group Stratified by Acute Myocardial Infarction (AMI) . Patients presenting with non-ST-elevation myocardial infarction (NSTEMI, n=63) and ST-elevation myocardial infarction (STEMI, n=92). The Shapiro-Wilk test assessed normality. In normally distributed data with continuous variables, a two-sided Student’s t-test was applied. In non-Gaussian distributed data, the Mann-Whitney test was used. Data are presented as the percent (%) of total or the 95% confidence interval, where applicable. Data are considered statistically significant if P≤0.05, denoted by **\***. BMI = body mass index, CHF = congestive heart failure, COPD = chronic obstructive pulmonary disease, CAD = coronary artery disease, CKD = chronic kidney disease, GRACE = Global Registry of Acute Coronary Events, MACE = major adverse cardiac events.

| **Internal Validation - Patient Demographics Stratified by Acute Myocardial Infarction (AMI)** |  |  |  |
| --- | --- | --- | --- |
| **Variable** | **NSTEMI (n=63)** | **STEMI (n=92)** | **P-Value** |
| Age (years) | 68.03 (66.48-69.58) | 65.47 (64.04-66.9) | 0.28 |
| Sex/Gender (Male) | 40 (63.49%) | 70 (76.09%) | 0.09 |
| Race/Ethnicity |  |  |  |
| Caucasian | 24 (38.1%) | 37 (40.22%) | 0.79 |
| Asian American | 8 (12.7%) | 22 (23.91%) | 0.08 |
| Hispanic American | 14 (22.22%) | 14 (15.22%) | 0.27 |
| Black/African American | 6 (9.52%) | 8 (8.7%) | 0.86 |
| BMI (kg/m<sup>2</sup>) | 27.82 (27.09-28.55) | 28.43 (27.42-29.44) | 0.67 |
| Systolic Blood Pressure (mmHg) | 143 (140-146) | 143 (140-147) | 0.96 |
| Diastolic Blood Pressure (mmHg) | 74 (72-75) | 80 (78-82) | 0.05 |
| Heart Rate (per minute) | 84 (81-86) | 85 (83-87) | 0.66 |
| Cardiac Arrest (at admission) | 0 (0%) | 4 (4.35%) | 0.09 |
| Troponin Elevation (at admission) | 63 (100%) | 89 (96.74%) | 0.15 |
| Smoking History |  |  |  |
| Current | 11 (17.46%) | 18 (19.57%) | 0.74 |
| Former | 18 (28.57%) | 22 (24.18%) | 0.54 |
| History of CHF | 1 (1.59%) | 19 (20.65%) | **\*0.0004** |
| History of COPD | 5 (7.94%) | 2 (2.17%) | 0.09 |
| History of CAD | 33 (52.38%) | 18 (19.57%) | **\*<0.0001** |
| History of CKD | 15 (23.81%) | 10 (10.87%) | **\*0.03** |
| History of Diabetes Mellitus | 35 (55.56%) | 39 (42.39%) | 0.11 |
| History of Hyperlipidemia | 38 (60.32%) | 51 (55.43%) | 0.55 |
| Prior Myocardial Infarction | 12 (19.05%) | 13 (14.29%) | 0.43 |
| Prior Percutaneous Intervention | 22 (34.92%) | 25 (27.17%) | 0.31 |
| Prior Coronary Artery Bypass Graft | 7 (11.11%) | 7 (7.61%) | 0.46 |
| Prior Stroke | 11 (17.46%) | 6 (6.52%) | **\*0.03** |
| GRACE Score | 107.92 (105.04-110.8) | 120.63 (116.97-124.28) | **\*0.02** |
| MACE at 30 Days | 6 (9.52%) | 13 (14.29%) | 0.38 |
| Cardiovascular Death - 1 year | 5 (8.06%) | 8 (8.7%) | 0.89 |
| All Cause Mortality - 1 year | 8 (12.70%) | 12 (13.04%) | 0.95 |

Echocardiographic functional features for patients in the internal validation study presenting with NSTEMI (n=63) and STEMI (n=92) were assessed (**Table 2**). Patients presenting with STEMI were more likely to have a LV ejection fraction (48% vs. 53%, P=0.0087) and left atrial end-systolic volume index (23 mL/m^2^ vs. 29 mL/m^2^, P=0.0024), compared to NSTEMI patients, respectively. Further the LV wall motion score index (2 vs. 1.7, P=0.0072) and LV global longitudinal strain (-11.86 vs. -14.1, P=0.0015) indicated greater wall motion abnormalities in STEMI compared to NSTEMI patients, respectively.

**Table 2:**
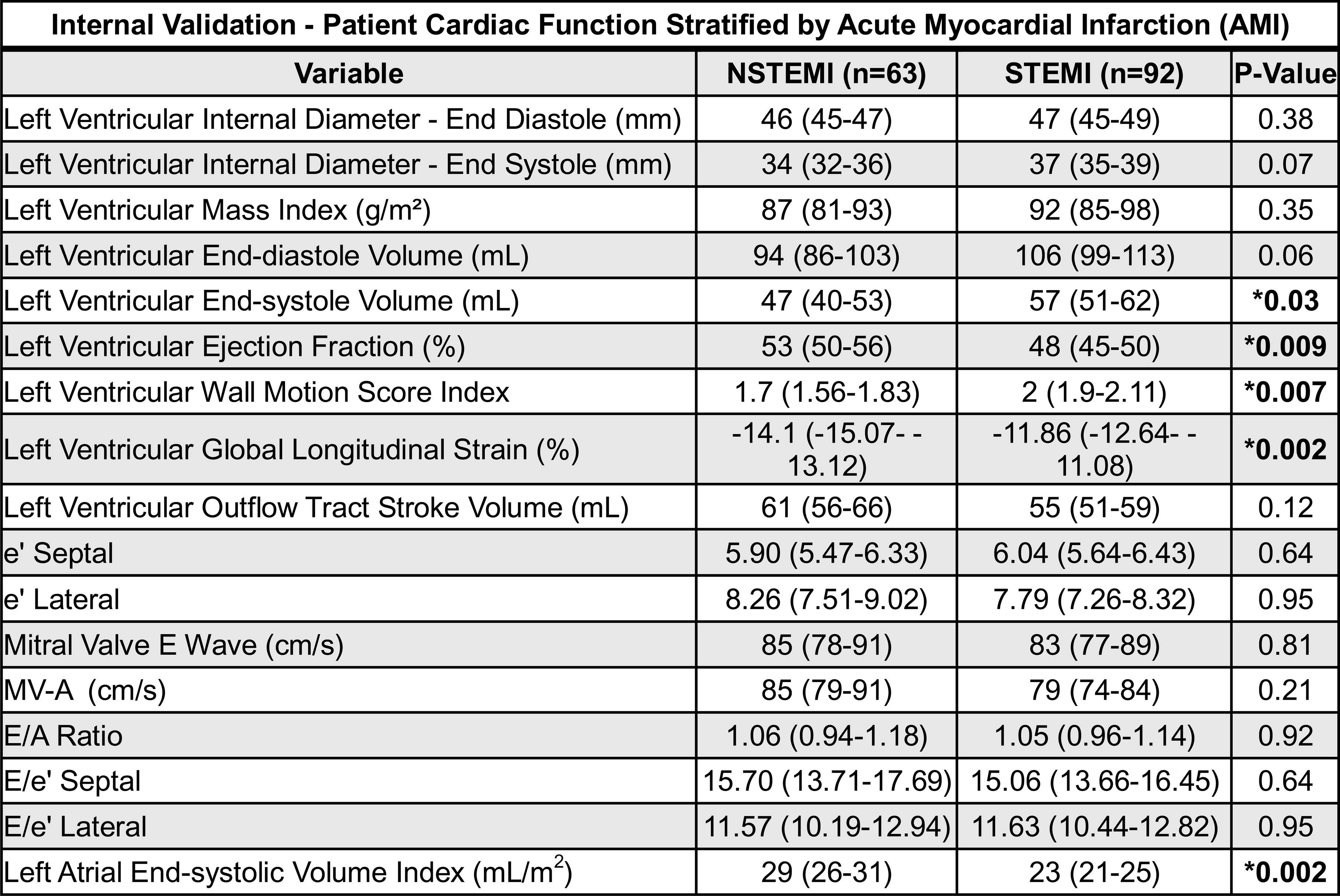
Patient Cardiac Function of the Internal Validation Group Stratified by Acute Myocardial Infarction (AMI) . Patients presenting with non-ST-elevation myocardial infarction (NSTEMI, n=63) and ST-elevation myocardial infarction (STEMI, n=92). The Shapiro-Wilk test assessed normality. In normally distributed data with continuous variables, a two-sided Student’s t-test was applied. In non-Gaussian distributed data, the Mann-Whitney test was used. Data are presented as the percent (%) of total or the 95% confidence interval, where applicable. Data are considered statistically significant if P≤0.05, denoted by **\***.

### Phenogroup Clustering through Topological Data Analysis (TDA)

Ultrasomics features were collected from the following echocardiography views: A4C, A3C, and A2C (**Figure 1A-B**). To understand if these features have value in predicting outcomes in patients presenting with AMI, ultrasomics features alone were evaluated using a TDA clustering algorithm that employed Mapper. Using the online tool TDAView, three phenogroups were identified: Cluster A (n=62), Cluster B (n=43), and Cluster C (n=50) (**Figure 2**). Of these phenogroups, Cluster A and Cluster B are illustrated to be more homogenous in their connectivity within groups, whereas Cluster C is illustrated to represent a more heterogenous compilation of patients.

**Figure 1:**
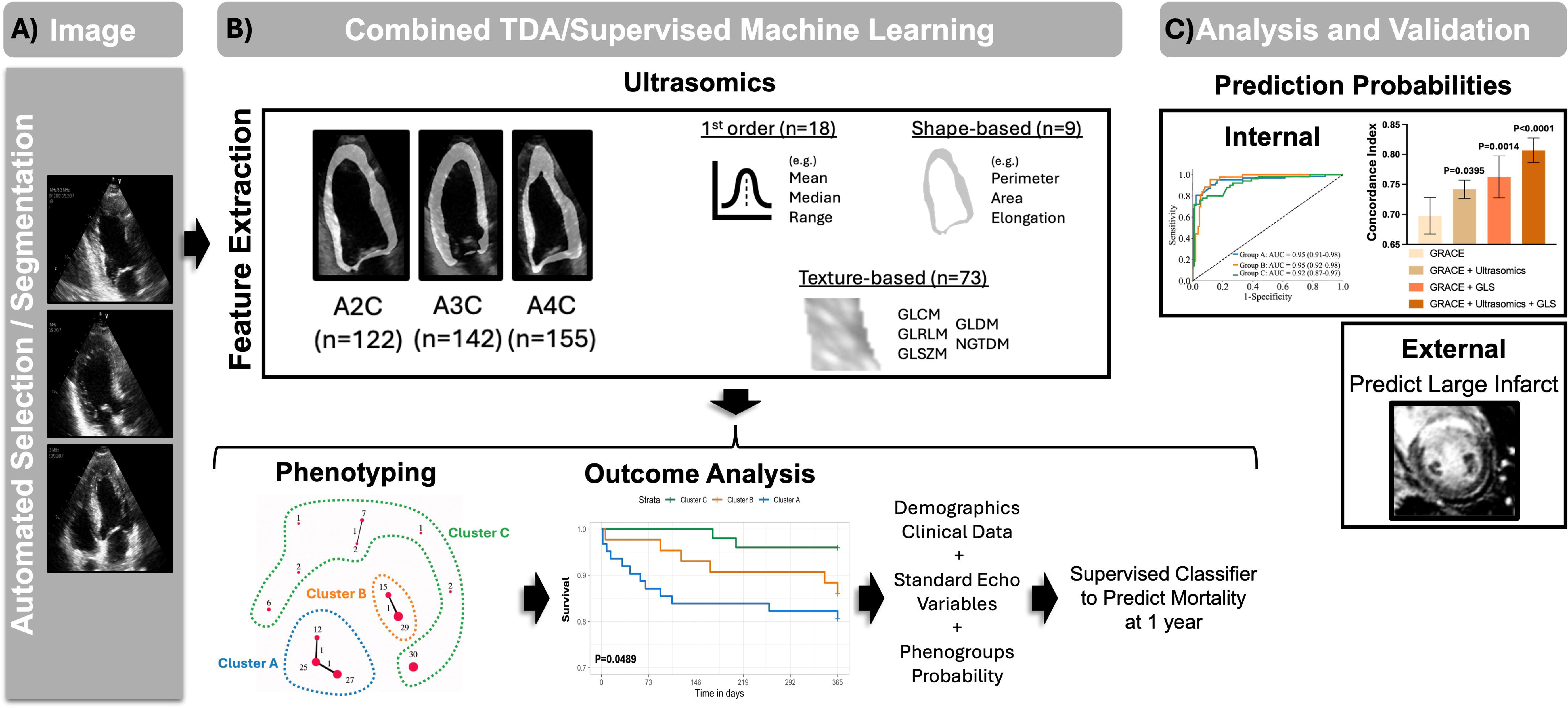
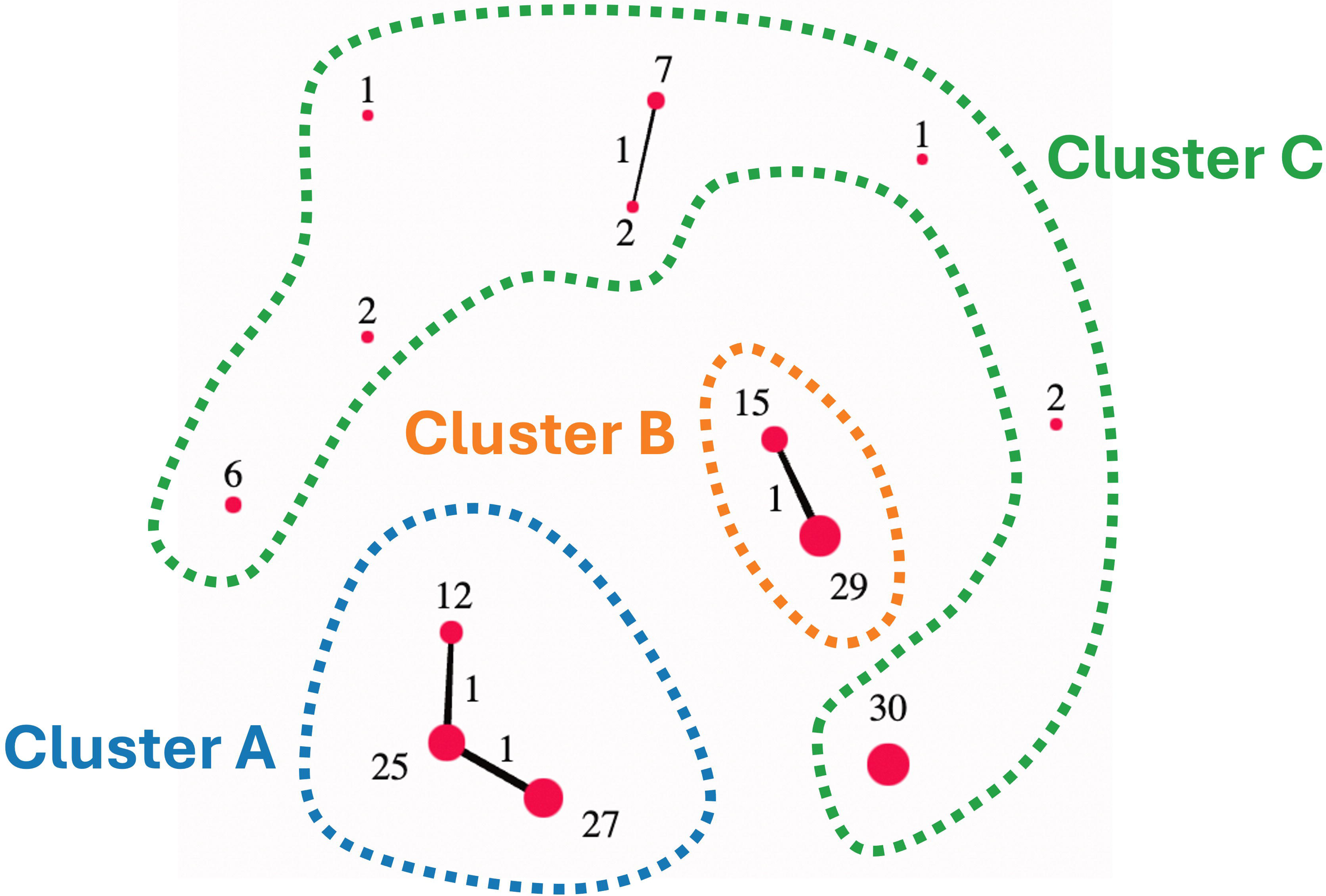
Study Design and Overview. (**A**) The internal validation patient cohort included patients with presenting with non-ST-elevation myocardial infarction (NSTEMI, n=63) and ST-elevation myocardial infarction (STEMI, n=92) who underwent echocardiography with views of the Apical 2-Chamber (A2C), Apical 3-Chamber (A3C), and Apical 4-Chamber (A4C). (**B**) Ultrasomics features were extracted using echocv and pyradiomics (v3.0.1). TDAView was used to cluster patients into three phenogroups: Cluster A, Cluster B, and Cluster C. The identified phenogroups were used to develop individual patient predicted probability of cluster assignment using a supervised machine learning classifier. (**C**) The generated probabilities from the supervised classifier were used to predict mortality and illustrate the incremental value of ultrasomics features over GRACE 2.0. Ultrasomics features were also extracted from the external validation group and applied to the supervised machine learning classifier to produce class labels (i.e., Cluster A, B, and C). The external validation phenogroups were used to predict findings on cardiac magnetic resonance, including acute infarct size.

**Figure 2:**
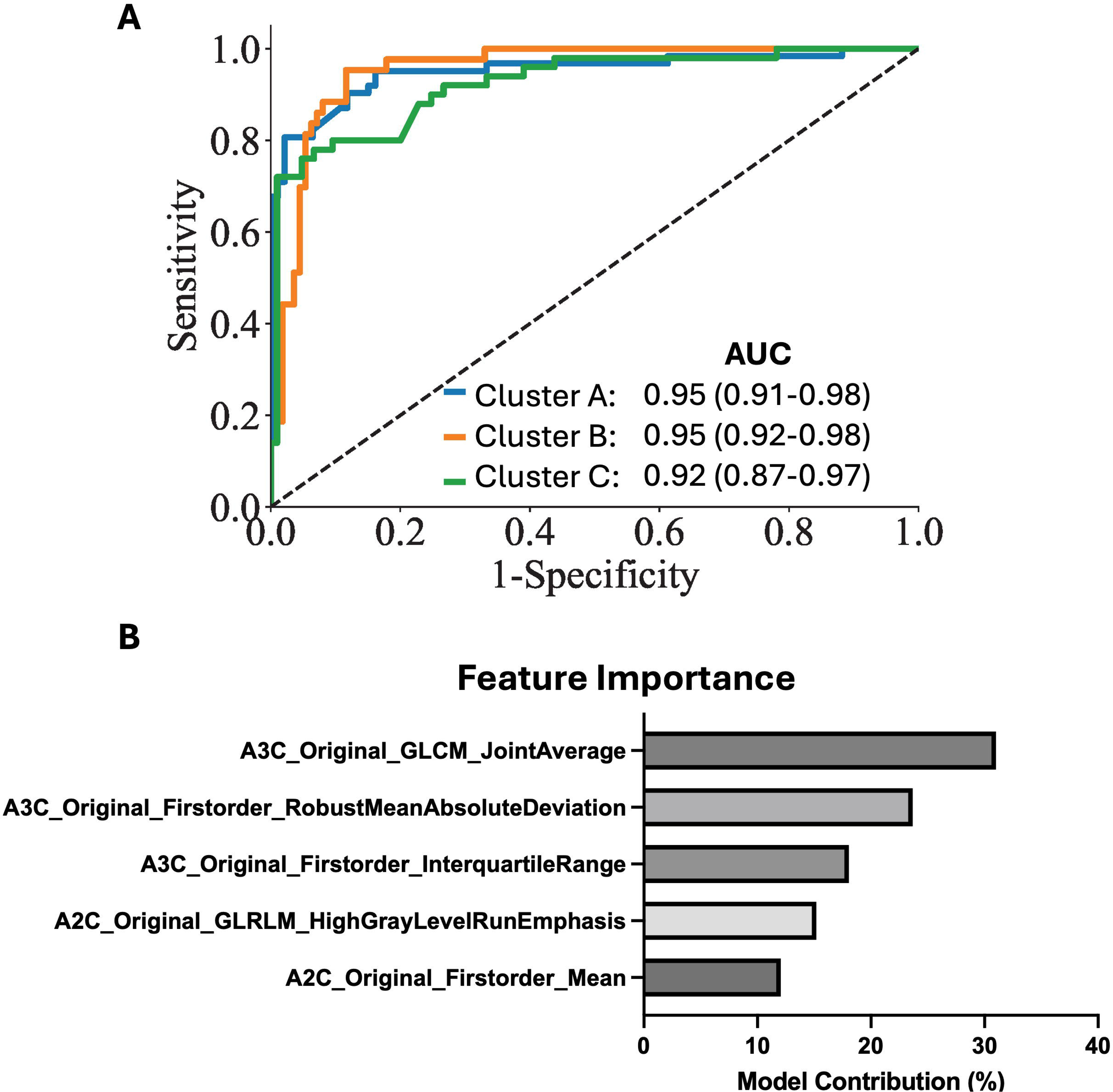
Topological Data Analysis (TDA) Clustering of Ultrasomics Features. Using TDAView a 1D Mapper filter was applied using Euclidean distance. Number of intervals was defined as 10, with 5 bins. To reduce the overlap between clusters, a 5% overlap was defined. Individual nodes are represented as red circles, with the number next to the node corresponding to the number of patients included in the node. Cluster A (n=62), Cluster B (n=43), and Cluster C (n=50).

Assessing the differences between these clusters, Cluster A contains more patients with a prior history of CHF (22.58% vs. 8%, P=0.0397), compared to Cluster C (**Table 3**). Further, the Cluster A phenogroup has a higher risk of all-cause mortality at 1 year (19.35% vs. 4%, P=0.0308), compared to Cluster C. The data in Table 2 highlight how the Cluster A represents a “high-risk” phenogroup, whereas Cluster B can be seen as “intermediate-risk” and Cluster C as “low-risk”. When assessing the echocardiographic functional parameters (**Table 4**), Cluster A had a reduced LV ejection fraction (45 vs. 53, P=0.0040) and LV global longitudinal strain (-11.88 vs. 13.87, P=0.0273) compared to Cluster C, respectively.

**Table 3:** Patient Demographics of the Internal Validation Group for Predicted Ultrasomics Phenogroups. Using only the ultrasomics features from the A4C, A3C, and A2C echocardiogram views, patients were clustered into phenogroups. Cluster A “high-risk” (n=62), Cluster B “intermediate-risk” (n=43), and Cluster C “low-risk” (n=50) using topological data analysis (TDA). A one-way analysis of variance (ANOVA) was applied for continuous variables and a Dunnett’s multiple comparisons test was used for multiple comparisons. For categorical data, a non-parametric Kruskal-Wallis test was applied with multiple comparisons testing. Data are presented as the percent (%) of total or the 95% confidence interval, where applicable. Data are considered statistically significant if P≤0.05, denoted by **\***. BMI = body mass index, CHF = congestive heart failure, COPD = chronic obstructive pulmonary disease, CAD = coronary artery disease, CKD = chronic kidney disease, STEMI = ST-elevation myocardial infarction, GRACE = Global Registry of Acute Coronary Events, MACE = major adverse cardiac events.

| Internal Validation - Patient Demographics in Predicted Ultrasomics Phenogroups |  |  |  |  |
| --- | --- | --- | --- | --- |
| Variable | Cluster A (High Risk)<br>(n=62) | Cluster B<br>(n=43) | Cluster C (Low Risk)<br>(n=50) | P-Value |
| Age (years) | 66.74 (62.98-70.51) | 66.88 (62.34-71.43) | 65.9 (62.03-69.77) | 0.94 |
| Sex/Gender (Male) | 44 (70.97%) | 31 (72.09%) | 35 (70%) | 0.98 |
| Race/Ethnicity |  |  |  |  |
| Caucasian | 24 (38.71%) | 16 (37.21%) | 21 (42%) | 0.89 |
| Asian American | 12 (19.35%) | 8 (18.6%) | 10 (20%) | 0.99 |
| Hispanic American | 9 (14.52%) | 9 (20.93%) | 10 (20%) | 0.64 |
| Black/African American | 5 (8.065%) | 4 (9.302%) | 5 (10%) | 0.94 |
| BMI (kg/m<sup>2</sup>) | 29.01 (26.08-31.93) | 28.9 (26.91-30.89) | 26.56 (24.68-28.43) | 0.28 |
| Systolic Blood Pressure (mmHg) | 140 (132-149) | 145 (135-155) | 145 (136-155) | 0.65 |
| Diastolic Blood Pressure (mmHg) | 78 (73-84) | 77 (71-84) | 76 (71-80) | 0.72 |
| Heart Rate (per minute) | 86 (81-92) | 85 (78-93) | 81 (76-87) | 0.47 |
| Cardiac Arrest (at admission) | 2 (3.226%) | 1 (2.326%) | 1 (2%) | 0.92 |
| Troponin Elevation (at admission) | 61 (98.39%) | 43 (100%) | 48 (96%) | 0.37 |
| STEMI (at admission) | 36 (58.06%) | 26 (60.47%) | 30 (60%) | 0.96 |
| Smoking History |  |  |  |  |
| Current | 16 (25.81%) | 11 (25.58%) | 13 (26.53%) | 0.99 |
| Former | 10 (16.13%) | 6 (13.95%) | 13 (26%) | 0.27 |
| History of CHF | 14 (22.58%)* | 2 (4.651%) | 4 (8%) | **\*0.01** |
| History of COPD | 1 (1.613%) | 4 (9.302%) | 2 (4%) | 0.17 |
| History of CAD | 21 (33.87%) | 18 (41.86%) | 12 (24%) | 0.19 |
| History of CKD | 10 (16.13%) | 5 (11.63%) | 10 (20%) | 0.55 |
| History of Diabetes Mellitus | 30 (48.39%) | 19 (44.19%) | 25 (50%) | 0.85 |
| History of Hyperlipidemia | 34 (54.84%) | 25 (58.14%) | 30 (60%) | 0.86 |
| Prior Myocardial Infarction | 8 (12.9%) | 10 (23.26%) | 7 (14.29%) | 0.34 |
| Prior Percutaneous Intervention | 5 (8.065%) | 5 (11.63%) | 4 (8%) | 0.67 |
| Prior Coronary Artery Bypass Graft | 21 (33.87%) | 13 (30.23%) | 13 (26%) | 0.79 |
| Prior Stroke | 6 (9.677%) | 4 (9.302%) | 7 (14%) | 0.71 |
| GRACE Score | 118.1 (109.1-127.2) | 114.5 (104.8-124.3) | 112.8 (103.9-121.8) | 0.69 |
| MACE at 30 Days | 7 (11.29%) | 5 (11.63%) | 7 (14.29%) | 0.88 |
| Cardiovascular Death - 1 year | 8 (13.11%) | 4 (9.302%) | 1 (2%) | 0.11 |
| All Cause Mortality - 1 year | 12 (19.35%)* | 6 (13.95%) | 2 (4%) | **\*0.04** |

**Table 4:**
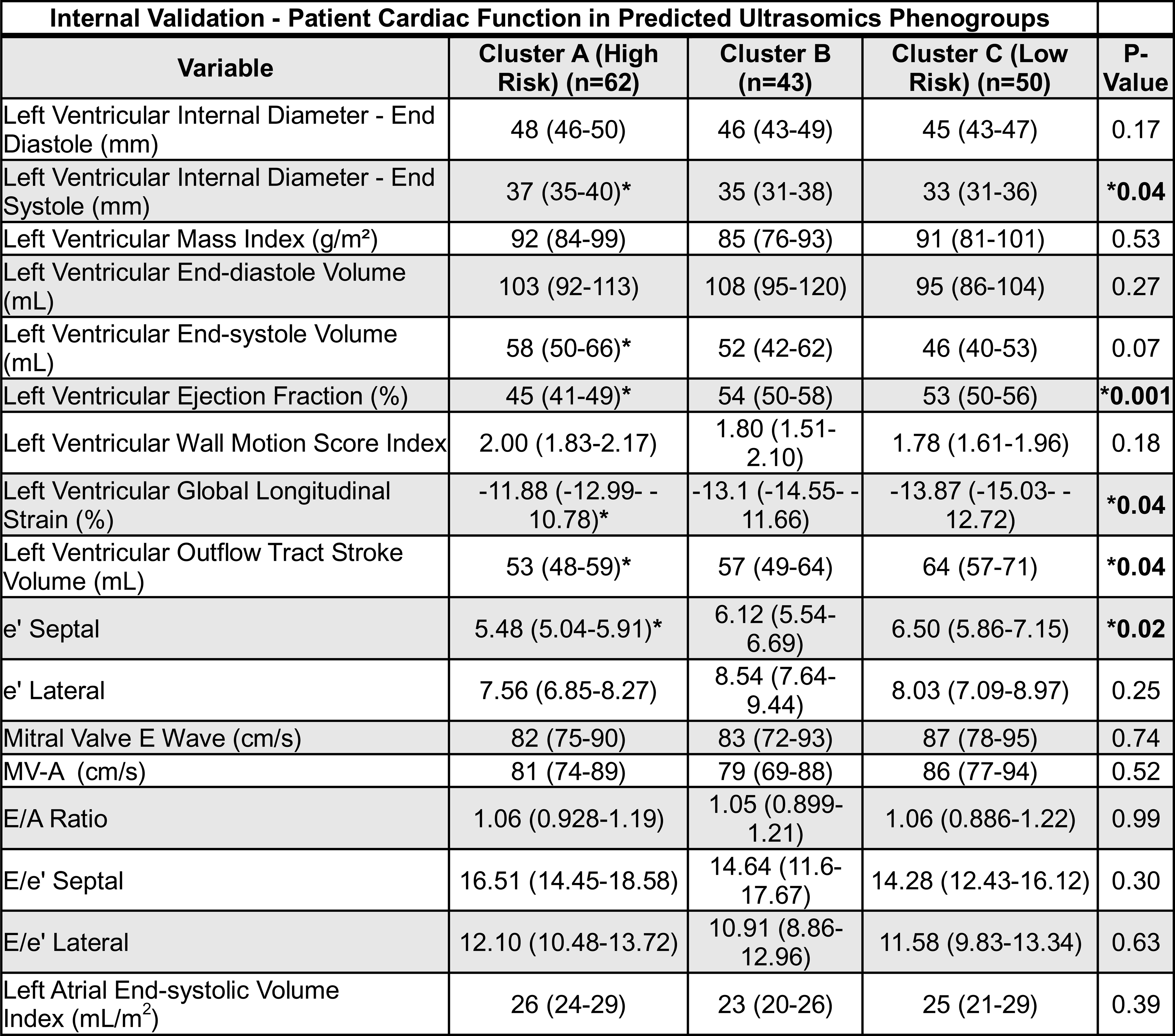
Patient Cardiac Function of the Internal Validation Group for Predicted Ultrasomics Phenogroups. Using only the ultrasomics features from the A4C, A3C, and A2C echocardiogram views, patients were clustered into phenogroups. Cluster A “high-risk” (n=62), Cluster B “intermediate-risk” (n=43), and Cluster C “low-risk” (n=50) using topological data analysis (TDA). A one-way analysis of variance (ANOVA) was applied for continuous variables and a Dunnett’s multiple comparisons test was used for multiple comparisons. For categorical data, a non-parametric Kruskal-Wallis test was applied with multiple comparisons testing. Data are presented as the percent (%) of total or the 95% confidence interval, where applicable. Data are considered statistically significant if P≤0.05, denoted by **\***.

### Supervised Machine Learning Classifier for Phenogroups

To establish individual patient-level probabilities to belong to specific phenogroups, a supervised machine learning classifier was developed using the online tool BigML with their OptiML application (i.e., mixed supervised model with 10-fold cross validation) on the internal validation patients. Using only ultrasomics features, the phenogroup labels were predicted for Cluster A (ROC AUC: 0.95), Cluster B (ROC AUC: 0.95), and Cluster C (ROC AUC: 0.92) (**Figure 3A**). When looking at the features contributing to the model, there was a mix of texture-based features and first order features (**Figure 3B**). Prediction probabilities were generated for the internal validation dataset based on the supervised classifier; these probabilities were used in subsequent analyses for risk prediction.

**Figure 3:**
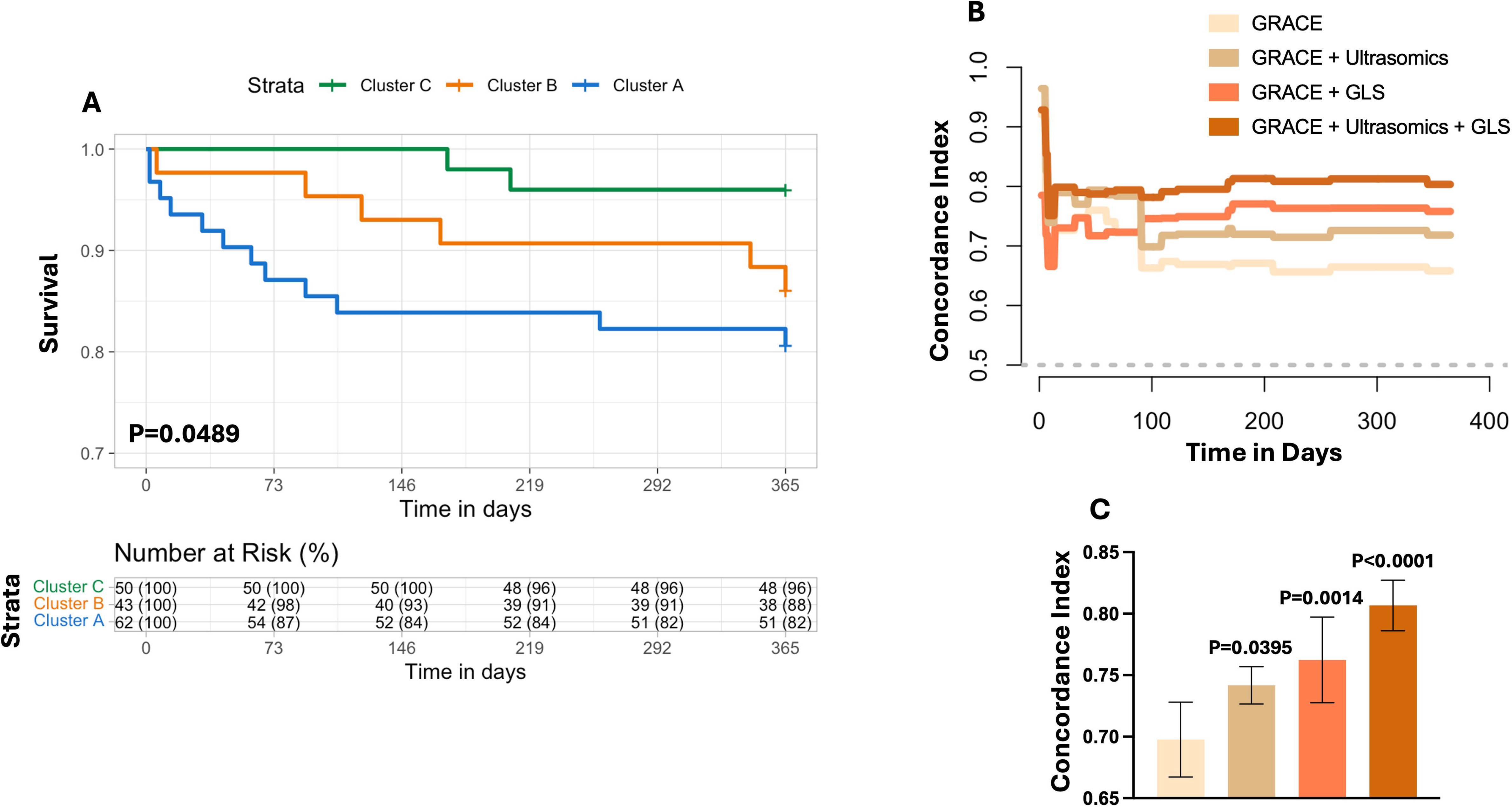
Supervised Machine Learning Classifier. (**A**) Ultrasomics features, as well as the class label for the topological data analysis (TDA)-defined phenogroups, were assessed using BigML and OptiML through 10-fold cross validation in the internal validation data. **(B**) The top five features contributing to model development for the supervised machine learning classifier.

### Outcome Prediction in the Internal and External Patient Groups

Using mortality at one year, survival analysis revealed that patients assigned to Cluster A had a significant increase in mortality compared to Cluster C (log rank, P=0.0489) (**Figure 4A**). We further wanted to further understand if the phenogroups, represented by changes in ultrasomics, had incremental value when predicting mortality. The concordance index was calculated for our four groups of variables: A) GRACE 2.0 score alone, B) GRACE + Cluster A, C) GRACE + LV global longitudinal strain, and D) using all three variables together (**Figure 4B**). We further illustrate that the use of ultrasomics alone (Concordance: 0.74 vs. 0.70, P=0.0395), and in combination with functional echocardiographic markers (Concordance: 0.81 vs. 0.70, P<0.0001), can increase prediction of all-cause mortality beyond that of the GRACE 2.0 score, respectively.

**Figure 4:**
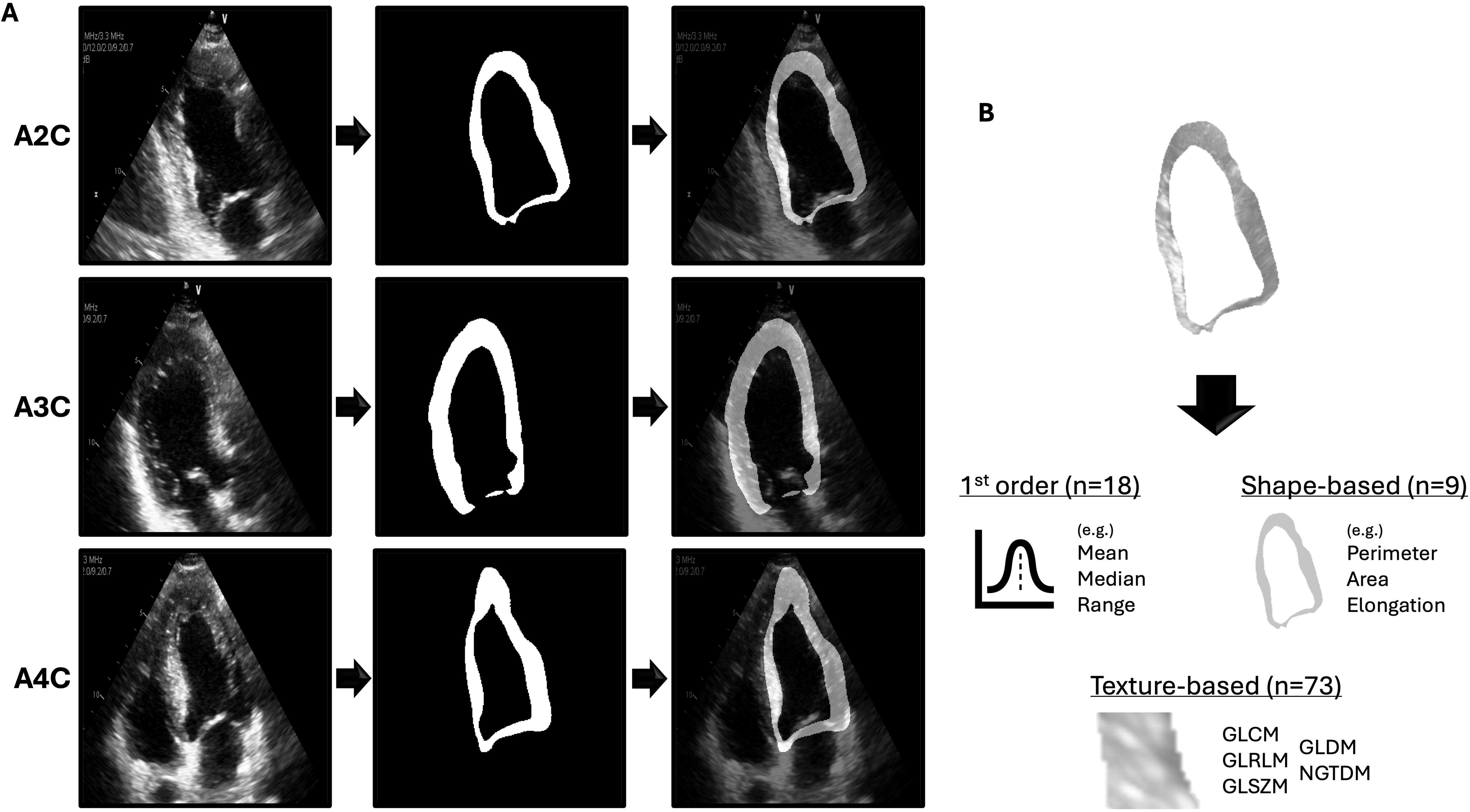
Performance of Phenogroups in Assessing All-Cause Mortality. (**A**) Kaplan Meyer curve and stratified risk categories for patients in phenogroups Cluster A, Cluster B, and Cluster C. (**B**) Time-to-event Concordance Index (C-Index) for groups A) GRACE 2.0 score alone, B) GRACE + Cluster A, C) GRACE + left ventricular global longitudinal strain (GLS), and D) using all three variables through CoxPH regression. (**C**) Incremental value of ultrasomics features (i.e., Cluster A) in predicting all-cause mortality. GRACE = Global Registry of Acute Coronary Events.

The developed supervised model was further applied to the external participants to assign phenogroup labels (i.e., Cluster A, B, and C). The batch prediction of the external dataset (n=42 presenting with STEMI) labeled participants into Cluster A (n=11), Cluster B (n=23), and Cluster C (n=8) (**Table 5**). Patients in Cluster A had a higher percentage of LV identified as “at risk” (60% vs. 37%, P=0.04) at 5 days post AMI, compared to Cluster C. Moreover, patients in the Cluster A phenogroup had a higher proportion of large infarcts (>10% of LV mass) at 30 days following AMI (0.91 vs. 0.25, P=0.007), when compared to Cluster C.

**Table 5:** Patient Demographics of the External Validation Group for Predicted Ultrasomics Phenogroups. Using the supervised machine learning classifier developed on the internal validation cohort, class labels were generated for the external hold out dataset (i.e., the prospective, multicenter, randomized DTU-STEMI pilot trial dataset) using batch prediction in BigML. Labels were applied based solely on ultrasomics features from the A4C, A3C, and A2C echocardiogram views. A one-way analysis of variance (ANOVA) was applied for continuous variables and a Dunnett’s multiple comparisons test was used for multiple comparisons. For categorical data, a non-parametric Kruskal-Wallis test was applied with multiple comparisons testing. Data are considered statistically significant if P≤0.05, denoted by **\***. BMI = body mass index, CHF = congestive heart failure, COPD = chronic obstructive pulmonary disease, CAD = coronary artery disease, CKD = chronic kidney disease, MACE = major adverse cardiac events, LV = left ventricular.

| External Validation - Patient Demographics in Predicted Ultrasonics Phenogroups |  |  |  |  |
| --- | --- | --- | --- | --- |
| Variable | Cluster A (High Risk)<br>(n=11) | Cluster B (n=23) | Cluster C (Low Risk)<br>(n=8) | P-Value |
| Age (years) | 56.82 (48.07-65.57) | 58.26 (53.72-62.8) | 62.88 (54.99-70.76) | 0.48 |
| Sex/Gender (Male) | 9 (81.82%) | 18 (78.26%) | 5 (62.5%) | 0.60 |
| Race/Ethnicity |  |  |  |  |
| Caucasian | 8 (72.73%) | 17 (73.91%) | 6 (75%) | 0.99 |
| Asian American | 1 (9.091%) | 3 (13.04%) | 0 (0%) | 0.58 |
| Hispanic American | 0 (0%) | 1 (4.545%) | 0 (0%) | 0.66 |
| Black/African American | 1 (9.09%) | 3 (13.04%) | 2 (25%) | 0.62 |
| BMI (kg/m<sup>2</sup>) | 30.08 (25.73-34.42) | 31.61 (27.35-35.88) | 25.61 (21.55-29.67) | 0.23 |
| Systolic Blood Pressure (mmHg) | 148 (129-167) | 158 (143-173) | 144 (132-157) | 0.44 |
| Diastolic Blood Pressure (mmHg) | 93 (81-104) | 91 (84-99) | 87 (75-99) | 0.71 |
| Heart Rate (per minute) | 85 (77-93) | 91 (81-101) | 81 (66-96) | 0.45 |
| Left Ventricular Ejection Fraction (%) | 36 (27-45) | 37 (31-43) | 44 (37-51) | 0.41 |
| Mitral Valve E Wave (cm/s) | 74 (58-91) | 77 (69-85) | 74 (60-89) | 0.93 |
| Mitral Valve A Wave (cm/s) | 72 (62-83) | 69 (59-78) | 74 (64-85) | 0.71 |
| E/A Ratio | 1.07 (0.78-1.37) | 1.20 (0.98-1.42) | 1.02 (0.79-1.24) | 0.55 |
| History of COPD | 0 (0%) | 0 (0%) | 0 (0%) | 0.99 |
| History of CAD | 1 (9.091%) | 1 (4.348%) | 1 (12.5%) | 0.73 |
| History of CKD | 0 (0%) | 0 (0%) | 0 (0%) | 0.99 |
| History of Diabetes Mellitus | 3 (27.27%) | 4 (17.39%) | 0 (0%) | 0.30 |
| History of Hyperlipidemia | 7 (63.64%) | 9 (39.13%) | 4 (50%) | 0.42 |
| Prior Stroke | 1 (9.091%) | 0 (0%) | 0 (0%) | 0.25 |
| MACE - 30 Days | 0 (0%) | 2 (8.696%) | 0 (0%) | 0.44 |
| Cardiovascular Death - 30 Days | 0 (0%) | 0 (0%) | 0 (0%) | 0.99 |
| All Cause Mortality - 30 Days | 0 (0%) | 0 (0%) | 0 (0%) | 0.99 |
| Infarct Size (%) of Area at Risk - 5 Days | 60 (52-68)* | 46 (37-56) | 37 (18-56) | 0.06 |
| Acute Volume of Infarct Size (mL) - 5 Days | 43 (32-54) | 31 (20-42) | 21 (-4.12-46) | 0.17 |
| Acute Infarct Size (%) of LV Mass - 5 Days | 23 (17-29) | 17 (11-23) | 12 (-0.53-25) | 0.24 |
| Acute Infarct Size >10% of LV Mass - 5 Days | 9 (82%) | 14 (61%) | 3 (38%) | 0.07 |
| Acute Volume of Infarct Size (mL) - 30 Days | 28 (21-36) | 23 (14-32) | 14 (-3.10-31) | 0.25 |
| Acute Infarct Size (%) of LV Mass - 30 Days | 18 (13-22) | 14 (8.73-19) | 9.23 (-1.31-20) | 0.27 |
| Acute Infarct Size >10% of LV Mass - 30 Days | 10 (91%) | 11 (48%) | 2 (25%) | **\*0.008** |

## Discussion

Properties of pathological changes within the myocardial microstructure influence ultrasound signal intensity distributions (31). Unlike information obtained indirectly (i.e., clinical risk factors, ECG, and biomarkers), specific analyzable trends in ultrasound texture information may have added insights into causal pathways that result in disease and clinical presentation. Integrating myocardial texture analysis (i.e., ultrasomics) with clinical data can provide a rich opportunity to develop machine learning models to predict adverse cardiac events following AMI. To this end we provide a proof-of-concept application of ultrasomics (i.e., cardiac ultrasound radiomics) in risk stratifying AMI patients. Three AMI phenogroups were identified according to ultrasound texture features with patients in phenogroup A having the worst prognosis. Phenogroup A showed incremental and independent information over GRACE 2.0 for predicting 1-year mortality after AMI. Using a cluster-then-predict framework we utilized an external hold out dataset for phenogroup prediction in which phenogroup A had large proportion of patients with moderate or large infarcts.

While classic supervised learning approaches require larger datasets, the cluster-then-predict methodology has the advantage of reducing bias, such as overfitting, when risk stratifying patients. Moreover this approach reduces prediction errors (37) and shows robust performance with echo-related data (38–41). Radiomics, deep learning features, 2D-echocardiography, demographic/clinical (e.g., age, sex, race, BSA, BMI, comorbidities, family history, etc.), laboratory, and biomarker data can further be added to incrementally increase the risk-stratification of these phenogroups. Our group has previously utilized TDA to create patient similarity networks to identify aortic stenosis (42), diastolic dysfunction (43–45), and heart failure (46,47). In aortic stenosis, by creating patient phenogroups for mild and severe aortic stenosis, the “high-risk” severe aortic stenosis phenogroup was associated with increased risk of balloon valvuloplasty, and valve replacement (42). Specifically, as shown in this study, the phenotypic groups from TDA (or unsupervised machine learning, PCA clustering, etc.) can serve as class labels for developing supervised algorithms. This technique, first clustering and then predicting using supervised machine-learning models, can result in stronger associations with clinical outcomes by increasing the number of events (i.e., phenogroup clusters) and reduce class imbalance.

Current risk stratification tools for AMI, such as the GRACE Score, reduce mortality rates compared to standard strategies (48,49) but, with the use of current AI applications, it is possible to characterize more patients at-risk for morbidity and mortality by combining information from clinical, laboratory, imaging, and other features. Risk stratification tools can be benchmarked using AUC and C-Index as metrics, with values ranging from 0.6-0.7 having limited clinical value, whereas those between 0.7-0.8, 0.8-0.9, and >0.9 considered to have fair, good, and excellent discrimination (50–52), respectively. The GRACE model has shown performances ranging from 0.65-0.8 (C-Index) (9), with our current study reporting a performance of 0.70, utilizing the GRACE 2.0 score. We also showed how the C-Index improved when using ultrasomics features (0.74) and in combination with LV functional parameters (0.81). As this is a pilot study, future work should harness these non-clinical markers (such as ultrasomics and LV functional information) in larger, multicenter studies to create new risk stratification tools for the prediction of AMI.

We note several limitations to the current investigation. 1) The cohort sizes in the internal and external validations sets are relatively small (n=155 and n=42, respectively). While this patient groups are small, we highlight how the cluster-then-predict methodology is better adapted to smaller datasets and can help provide a framework for other investigations where small cohort sizes are present (i.e., rare diseases, underrepresented minorities, limited resources for data collection, etc.). 2) The outcome of interest, all-cause mortality at 1 year, was only represented in 20 of 155 patients. Because of the low number of events, we used univariate analysis to screen for features to provide in the adjusted model while avoid issues with overfitting in the survival model. Nevertheless, we noted the incremental value of radiomics over conventional scores like Grace 2.0 and several echocardiographic parameters like ejection fraction, LV end-systolic volume and global longitudinal strain. Future work with larger sample size and a greater number of events would allow develop of robust multivariable models using radiomics, clinical and conventional echocardiographic features. 3) The use of TDA, and other unsupervised learning approaches, can be subjective in the number of clusters defined. In the current study, we highlight three unique phenogroups. While we could have altered the parameters to include more or less numbers of phenogroups, the main constraint on the Mapper algorithm that we wanted to maintain was a low percent overlap between groups (i.e., reducing the similarities of phenogroups and ultimately providing clearer boundaries between those with “high” and “low” risk).

In summary, we utilize an echocardiography-derived approach to measure ultrasomics and identify phenogroups among patients presenting with AMI. Through TDA, three distinct phenogroups (Clusters A, B, and C) were delineated, with Cluster A representing a “high-risk” group, Cluster B an “intermediate-risk” group, and Cluster C a “low-risk” group. These phenogroups demonstrated significant differences in clinical outcomes, particularly in terms of all-cause mortality at 1 year. Logistic regression and supervised machine learning further validate the predictive power of these phenogroups, showing their potential utility in clinical risk stratification. Moreover, application of the developed model to an external dataset highlighted the robustness of these phenogroups in predicting cardiac magnetic resonance (CMR) findings such as infarct size, providing valuable insights for personalized patient management and prognostication in AMI.

## Acknowledgments

None

## Author Contributions

