## Supplemental Figure S1 for "Ultrasonic Texture Analysis for Acute Myocardial Infarction Risk Stratification: A Pilot Study"

*Running Title: Ultrasonic Texture and Myocardial Infarction Risk Stratification*

*****Provided equal contribution to the work

**Corresponding Author**

Partho P. Sengupta, MD, DM, FACC, FASE

Rutgers Robert Wood Johnson Medical School,

Division of Cardiovascular Disease and Hypertension,

125 Patterson St, New Brunswick, NJ – 08901

**Figure S1**

**Figure S1**: **Illustration of Left Ventricular Ultrasomics in Apical 2-Chamber (A2C), Apical 3-Chamber (A3C), and Apical 4-Chamber (A4C).** (**A**) Representative images for region of interest (ROI) placement using semantic segmentation through echocv. (**B**) Example of ultrasomics features extracted from the Python package pyradiomics (v3.0.1), including 1^st^ order (n=18), shape-based (n=9), and texture-based (n=73) features. GLCM = gray-level cooccurrence matrix, GLDM = gray-level difference matrix, NGTDM = Neighborhood gray-tone difference matrix, GLRLM = gray-level run-length, GLSZM = gray-level size zone matrix.
